# Neonatal brain perivascular space volume as a predictor of neurodevelopmental outcomes at 24 months

**DOI:** 10.1101/2025.05.26.25328343

**Authors:** Arum Choi, Dayeon Bak, Ah-Ra Cho, Hosna Asma-ull, Yoonho Nam, Hyun Gi Kim

## Abstract

**Introduction:** Perivascular space (PVS) has recently gained attention as a neurological indicator associated with early brain development in infants. However, there is still limited research on the relationship between PVS volume and neurodevelopmental outcome of infants. Therefore, this study aimed to investigate the association between PVS volume of neonates at term-equivalent age and their neurodevelopmental outcome at 24 months.

**Methods:** This study retrospectively reviewed neonates born between 2019 and 2022 at a single institution who underwent brain MRI at term-equivalent age and had neurodevelopmental assessment at 24 months using Bayley-III. Neonates with structural abnormalities on brain MRI due to clinical risk factors such as congenital anomalies or infections were excluded. PVS volume of the brain MRI were automatically extracted and relationship between the volume and neurodevelopmental scores on three domains (cognitive, language, and motor) were assessed using multiple linear regression. We conducted Wilcoxon rank-sum tests to compare PVS volumes between infants with normal and delayed development group, defined as a composite score below 85 in each outcome domain.

**Results:** A total of 14 neonates were included (median gestational age, 246 [243–254] days; birth weight, 3 [2– 3] kg). Result of multiple linear regression analysis showed significant negative associations between PVS volume and cognitive (β = -0.70, *P* = 0.042), language (β = -0.69, *P* = 0.01), and motor development (β = -0.71, *P* = 0.017) after adjusting for corrected gestational age. Infants with developmental delays demonstrated significantly higher median PVS volumes compared to those with normal development for language (38.4 vs 18.5 mm^3^, *P* = 0.02) and motor domains (51.2 vs 26.6 mm^3^, *P* = 0.02).

**Conclusion:** Increased neonatal brain PVS volume was associated with lower developmental scores in cognitive, language, and motor domains at 24 months. Additionally, greater PVS volume independently predicted language developmental delay, suggesting that neonatal PVS volume may serve as an independent predictor of neurodevelopmental outcomes.

## Introduction

[Neonatal brain MRI and neurodevelopmental outcome] Neonatal brain magnetic resonance imaging (MRI) is recognized as an important tool for predicting neurodevelopmental outcomes (Woodward et al., 2006; Anderson et al., 2015), especially in infants at high risk for neurodevelopmental disorders, such as premature infants (Schmidbauer et al., 2024). The presence of MRI abnormalities, most notably moderate-to-severe white matter injury (Woodward et al., 2006), deep gray matter abnormalities (Woodward et al., 2006), cerebellar lesions (Park et al., 2014), and maturational delays(Slaughter et al., 2016), has strong and specific associations with subsequent cognitive, motor, and sensory impairments. Brain MRI performed in the neonatal period, especially at term-equivalent age, serves as a critical noninvasive biomarker for assessing long-term neurodevelopmental outcome in preterm and high-risk infants(Yin et al., 2021).

[MRI PVS volume as glymphatic function marker] Recent studies have focused on the MRI-visible perivascular space (PVS) volume, which may reflect the function of the glymphatic system, the brain’s waste removal system (Kim et al., 2023; Naganawa et al., 2024). Changes in PVS volume in newborns, particularly in the basal ganglia, have been associated with maturation and prematurity, which may potentially be associated with neurodevelopmental outcomes. For example, one study found that newborns with hypoxic-ischemic brain injury (HII) had smaller basal ganglia PVS volumes compared to uninjured newborns, suggesting that PVS volume assessment may serve as a biomarker of glymphatic function changes (Choi et al., 2025). Another study reported that enlarged PVS measured at 6, 12, and 24 months was associated with a diagnosis of autism at 24 months (Garic et al., 2023).

[Glymphatic function and neurodevelopment disorders] The glymphatic system facilitates the elimination of metabolic byproducts from cerebral parenchymal tissue via convective transport mechanisms involving cerebrospinal fluid-interstitial fluid exchange (Gao et al., 2023). During the neonatal period, glymphatic system maturation plays an essential role in normal brain structure and function development. When this system is underdeveloped or dysfunctional, the risk of neurodevelopmental disorders may increase (Lin et al., 2024; Agarwal et al., 2025). This vulnerability is particularly pronounced in premature infants, where the developing glymphatic system’s immaturity potentially contributes to increased risk of neurodevelopmental disorders such as autism spectrum disorder (ASD) (Li et al., 2022) or Attention-Deficit Hyperactivity Disorder (ADHD) (Chen et al., 2024).

[Objective] However, no studies to date have investigated whether neonatal PVS volume can predict later developmental outcomes. Therefore, the objective of this study was to examine the association between PVS volume on brain MRI at term-equivalent age and neurodevelopmental outcomes at 24 months.

## Materials and Methods

### Study sample

This study was a retrospective study at a single center. We included neonates born between 2019 and 2022 who underwent brain MRI at term-equivalent age and had neurodevelopmental assessment completed at 24 months using the Bayley Scales of Infant and Toddler Development, Third Edition (Bayley-III). We excluded neonates with structural abnormalities on brain MRI due to clinical risk factors such as congenital anomalies or infections. We also excluded neonates who were lost to follow-up before 24-month assessment. We collected demographic and clinical data from electronic medical records (EMR), including gestational age at birth (GA), birth weight, sex, mode of delivery and Apgar scores at 1 and 5 minutes.

### MRI acquisition

Brain MRI scans were acquired using a 3T scanner (Magnetom Vida, Siemens Healthineers) equipped with a specialized 64-channel receiver coil designed for neonatal imaging. All scans were acquired during natural sleep without sedation. The imaging protocol included three-dimensional T1-weighted Magnetization-Prepared Rapid Acquisition with Gradient Echo (MPRAGE) sequences, three-dimensional T2-weighted Sampling Perfection with Application-Optimized Contrasts using Different Flip Angle Evolution (SPACE) sequences, diffusion tensor imaging (DTI), and susceptibility-weighted imaging (SWI).

### PVS volume extraction

We initially segmented brain region automatically using Infant FreeSurfer (Zollei et al., 2020) followed by Frangi filtering to enhance enlarged PVS voxels (Frangi et al., 1998; Kim et al., 2023). Multiple thresholding techniques were then implemented, incorporating standard deviation multiples added to mean intensity values within the BG region (caudate, putamen, and pallidum) to isolate high-intensity voxels. A trained researcher subsequently conducted manual segmentation refinement under supervision of an experienced pediatric neuroradiologist with 15 years of expertise to enhance quantification precision. The refinement protocol encompassed removal of false-positive PVS identifications, correction of false-negative PVS detections, and threshold optimization to enhance boundary definition between BG and PVS regions. The BG-PVS volume was obtained by quantifying the total number of segmented voxels per unit volume. All manual corrections were conducted using ITK-SNAP version 4.2.0 (Yushkevich et al., 2016), with remaining computational tasks performed in Python version 3.8 (Python Software Foundation).

### Neurodevelopment outcome assessments

Neurodevelopmental outcomes were collected through EMR review. Results of neurodevelopmental assessments administered using the Bayley-III at 24 months were retrospectively reviewed. The Bayley-III assesses neurodevelopmental function in three major domains: cognitive development (sensorimotor skills, exploration and manipulation, object relatedness, concept formation, memory), language development (receptive and expressive communication skills), and motor development (fine and gross motor skills) (Johnson et al., 2014; Ahn and Kim, 2017). Based on composite scores extracted from the EMR, normal neurodevelopment was classified as a composite score ≥85 in each domain, and delayed neurodevelopment as a composite score <85 (Johnson et al., 2014; Spencer-Smith et al., 2015).

### Statistics

We evaluated continuous variables for normality using the Shapiro-Wilk test and presented them as median [interquartile range (IQR)], while categorical variables were described as frequency (percentage). For the primary analysis, we used multiple linear regression models to examine the relationship between BG-PVS volume and Bayley-III composite scores in each domain: cognitive, language, and motor. Models were adjusted for corrected GA (CGA) at the time of MRI scans. Due to the small sample size, only one confounding variable was included in the model, selected based on clinical relevance and univariate association with the outcome (*P* < 0.2). For the secondary analysis, we performed Wilcoxon rank-sum tests to compare BG-PVS volumes between infants with normal (≥85) and delayed (<85) development in each outcome domain. The statistical significance level was set at *P* < 0.05. All analyses were performed using R version 4.4.1.

## Results

### Study sample

The study included 14 infants with complete data for analysis (Table 1). Infants were born at a median GA of 246 [243–254] days and a median CGA of 263 [259–275] days, with birth weights median 3 [2–3] kg. The median Apgar scores were 6 at 1 minute and 8 at 5 minutes. At 24-month neurodevelopmental assessment by Bayley-III, median composite scores were 98 [91–105] for cognitive development, 79 [77–93] for language development, and 100 [92–114] for motor development. PVS volume measurements ranged widely among participants (29.18 [22.27–41.09] mm^3^).

**Table 1.** Demographic and Clinical Characteristics of Study Samples.

|  | Median [IQR] |
| --- | --- |
| **Birth Characteristics** |  |
| Gestational age (days) | 246 [243–254] |
| Corrected gestational age (days) | 263 [259–275] |
| Birth weight (kg) | 3 [2–3] |
| **Neonatal Assessment** |  |
| Apgar score 1min | 6 [5–8] |
| Apgar score 5min | 8 [8–9] |
| **24-Month Developmental Outcomes** |  |
| Cognitive composite score | 98 [92–105] |
| Language composite score | 79 [77–93] |
| Motor composite score | 100 [92–111] |
| **MRI Biomarker** |  |
| PVS volume (mm<sup>3</sup>) | 29.18 [22.27–41.09] |
| PVS, perivascular space; IQR, interquartile range |  |

### Relationship between outcome and PVS volume

Based on univariate screening criteria (*P* < 0.2), CGA was included as a confounding variable in all final models. The multiple linear regression analyses revealed significant negative associations between PVS volume at term-equivalent age and neurodevelopmental composite scores at 24 months across all three developmental domains (Table 2). For cognitive development, higher PVS volume was associated with lower cognitive composite scores (β = -0.70, *P* = 0.04). Similarly, higher PVS volume was significantly associated with lower language composite scores (β = -0.69, *P* = 0.01). For motor development, higher PVS volume was associated with lower motor composite scores (β = -0.71, *P* = 0.02).

**Table 2.** Multiple linear regression analysis examining associations between PVS volume and 24-month neurodevelopmental outcomes, adjusted for CGA.

| | $\beta$ | SE | t value | P-value |
| --- | --- | --- | --- | --- |
| **Cognitive Development** |  |  |  |  |
| PVS volume | -0.70 | 0.30 | -2.30 | 0.04 |
| CGA | -0.07 | 0.26 | -0.29 | 0.78 |
| **Language Development** |  |  |  |  |
| PVS volume | -0.69 | 0.24 | -2.93 | 0.01 |
| CGA | 0.13 | 0.20 | 0.64 | 0.54 |
| **Motor Development** |  |  |  |  |
| PVS volume | -0.71 | 0.25 | -2.80 | 0.02 |
| CGA | -0.50 | 0.22 | -2.28 | 0.04 |

P-values from multiple linear regression. PVS: perivascular space; CGA: corrected gestational age; $\beta$ : beta coefficient; SE: standard error

### PVS volume as a predictor for neurodevelopmental outcome

The distribution of PVS volumes between infants with normal versus delayed development across three developmental domains is summarized in Table 3. Infants with delayed development consistently showed higher PVS volumes compared to those with normal development. For cognitive development, infants with delays (n=3) had significantly higher median PVS volumes compared to those with normal development (n=11) (45.1 [42.0–48.0] mm^3^ vs 26.0 [20.0–36.0] mm^3^, *P* = 0.06). The association was statistically significant for language development, where infants with language delays (n=9) demonstrated higher PVS volumes than those with normal language development (n=5) (38.4 [28.0–45.0] mm^3^ vs 18.5 [17.0–24.0] mm^3^, *P* = 0.02). Similarly, for motor development, the two infants with motor delays showed higher PVS volumes compared to those with normal motor development (n=12) (51.2 [50.0–52.0] mm^3^ vs 26.6 [21.0– 34.0] mm^3^, *P* = 0.02).

**Table 3.** Comparison of PVS volumes between normal and delayed development groups.

|  | Normal Development |  | Delayed Development |  | P-value |
| --- | --- | --- | --- | --- | --- |
|  | N | Median [IQR] | N | Median [IQR] |  |
| **Cognitive Development** | 11 | 26 [20–36] | 3 | 45 [42–48] | 0.06 |
| **Language Development** | 5 | 19 [17–24] | 9 | 38 [28–45] | 0.02 |
| **Motor Development** | 12 | 27 [21–34] | 2 | 51 [50–52] | 0.02 |

P-values are from Wilcoxon rank-sum tests. PVS: perivascular space; CGA: corrected gestational age; IQR, interquartile range

## Data Availability

The datasets used and analyzed during the current study are available from the corresponding author on reasonable request, subject to institutional review board approval and ethical considerations.

